# The mental health impact of COVID-19 racial and ethnic discrimination against Asian American and Pacific Islanders

**DOI:** 10.1101/2021.06.06.21258177

**Authors:** Sasha Zhou, Rachel Banawa, Hans Oh

## Abstract

Hate crimes against Asian American/ Pacific Islanders (AAPIs) have surged in the United States during the COVID-19 pandemic to alarming new levels. We analyzed data from the Healthy Minds Study, and found that COVID-19 related racial/ethnic discrimination was associated with greater odds of having depression, anxiety, non-suicidal self-injury, binge drinking, and suicidal ideation among AAPI university students (N=1697). Findings suggest that the COVID-19 pandemic precipitated discrimination, which has been linked to mental health problems, calling for more preventive interventions to address the AAPI population, especially given their low rates of formal treatment utilization.

## INTRODUCTION

Racism against Asian American Pacific Islanders (AAPI) is not a new phenomenon in the United States, but reports of discrimination and hate crimes against this community have surged to new heights during the COVID-19 pandemic. According to the Pew Research Center^1^, about 40% of Asian American adults reported that other people were visibly uncomfortable around them since the start of the pandemic. According to the Center for the Study of Hate and Extremism^2^, anti-Asian hate crimes increased by almost 150% across 16 of the country’s largest cities in the year 2020. And between March 19, 2020 to February 28, 2021, the Stop AAPI Hate reporting center documented 3,795 hate incidents, ranging from online harassment to physical assault^3^. The list of hate crimes is harrowing and continues to grow^4^; we should note that many hate crimes go unreported. Much of this racism has been fueled by a xenophobic narrative that AAPI’s are somehow responsible for the COVID-19 pandemic, underscoring a long-held view that AAPIs are perpetual foreigners who do not belong in the country^5^. This racialization of COVID-19 has the potential to produce long-lasting effects on attitudes towards AAPIs, which is alarming since a substantial body of research has linked racial discrimination to adverse mental health outcomes as well as lower use of formal psychiatric treatment^6^. In this study, we analyzed a sample of AAPI university students from across the country to examine the associations between COVID-19 related racial/ethnic discrimination and mental health outcomes during the pandemic.

## METHODS

We analyzed data the 2020 Healthy Minds Study (HMS), which is a cross-sectional, web-based survey examining mental health and related factors in students enrolled at one of 28 universities. The HMS is designed to protect the privacy and confidentiality of participants, and has been approved by the Health Sciences and Behavioral Sciences Institutional Review Board at University of Michigan. To further protect respondent privacy, the study is covered by a Certificate of Confidentiality from the National Institutes of Health. The study survey was administered between September through December of 2020. We focused on students who self-identified as AAPI (N=1697). The mean age of this AAPI sample was 23.78 years old (95%CI: 23.15-24.41), and the majority was cis-gendered women (67.88%; n=1152).

Respondents were asked a single binary item (yes/no): *As a result of the COVID-19 pandemic, have you experienced any discriminatory or hostile behavior due to your race/ethnicity (or what someone thought was your race/ethnicity)*? We examined this question in relation to several mental health outcomes: depression, anxiety, binge drinking, non-suicidal self-injury, and suicidal ideation.

## RESULTS

Among the AAPI students, over a quarter reported experiencing COVID-19 related racial/ethnic discrimination. Over two-thirds of respondents who endorsed this item met the criteria for at least one clinically significant mental health condition. Using multivariable logistic regression models, we found that COVID-19 related racial/ethnic discrimination was associated with greater odds of having moderately severe or severe depression, moderate to severe anxiety, any binge drinking over the past two weeks, non-suicidal self-injury, and suicidal ideation, adjusting for age and gender.

## DISCUSSION

These findings should be interpreted bearing in mind that racial/ethnic discrimination was self-reported, which is prone to both under- and over-reporting^7^. Moreover, the study used a non-probability sampling strategy that yielded a response rate of 14%, which is admittedly low but common for these types of online surveys^8^. We did however use sample probability weights to adjust for non-response using the following administrative data on full student populations: gender, race/ethnicity, academic level, and Grade Point Average. Still, it remains to be seen whether these associations are generalizable to the larger AAPI population and global Asian population; it is possible that the associations may be even stronger outside of the university context, especially among immigrants with limited English proficiency.

Historically, less than a third of AAPI students with a clinically significant mental health condition are engaged in mental health treatment, which is the lower than other racial groups^8^. Preventive interventions are needed to eliminate this treatment gap. Undoubtedly, anti-Asian discrimination and hate crimes continue to devastate individuals and communities across the world, and so as AAPI researchers, we urge our colleagues and institutions to speak out publicly against this hatred, to design interventions that mitigate the pernicious effects of racism on population health, and to call for the removal of barriers that prevent racial and ethnic minorities from accessing appropriate mental health treatment.

**Table 1.** Associations between COVID-19 related racial/ethnic discrimination and mental health outcomes among American/Pacific Islander students from the Healthy Minds Study, September – December 2020.

|  | Descriptive statistics |  |  |  | Multivariable logistic regression models |
| --- | --- | --- | --- | --- | --- |
|  | Total<br>N=1697 | No discrimination<br>N=1262 (74.37%) | Discrimination<br>N=435 (25.63%) | P-value | aOR [95% CI] |
| **Depression** |  |  |  |  |  |
| No | 1395 (83.88%) | 1073 (86.95%) | 322 (75.06%) | <0.001 | 1.00 |
| Yes | 268 (16.12%) | 161 (13.05%) | 107 (24.94%) |  | 2.02 [1.55-2.62] |
| **Anxiety** |  |  |  |  |  |
| No | 948 (76.58%) | 1203 (72.30%) | 255 (59.86%) | <0.001 | 1.00 |
| Yes | 290 (23.42%) | 461 (27.70%) | 171 (40.14%) |  | 1.97 [1.58-2.44] |
| **Binge drinking** |  |  |  |  |  |
| No | 1372 (80.85%) | 1039 (82.33%) | 333 (76.55%) | <0.001 | 1.00 |
| Yes | 325 (19.15%) | 223 (17.67%) | 102 (23.45%) |  | 1.46 [1.17-1.83] |
| **Non-suicidal self-injury** |  |  |  |  |  |
| No | 1364 (81.68%) | 1050 (84.54%) | 314 (73.36%) | <0.001 | 1.00 |
| Yes | 306 (18.32%) | 192 (15.46%) | 114 (26.64%) |  | 1.78 [1.40-2.27] |
| **Suicidal ideation** |  |  |  |  |  |
| No | 1545 (91.26%) | 1172 (93.09%) | 373 (85.94%) | <0.001 | 1.00 |
| Yes | 148 (8.74%) | 87 (6.91%) | 61 (14.06%) |  | 2.02 [1.43-2.85] |

P-values reflect t-test for continuous variables and Chi<sup>2</sup> test for binary/categorical variables.
(1) Depression was measured using the Patient Health Questionnaire 9, which is a scale ranging from 0-27. We dichotomized the scale to reflect individuals who reported a score higher than 15, indicating moderately severe to severe depression.
(2) Anxiety was measured using the Generalized Anxiety Disorder 7, which is a scale ranging from 0-21. We dichotomized this scale to reflect individuals who reported a score of higher than 10, indicating moderate to severe anxiety.
(3) Binge drinking was assessed using the item: Over the past 2 weeks, did you have 4 [if female] / 5 [if male] or more alcoholic drinks in a row?
(4) Self-injury was assessed using the item: In the past year, have you ever done any of the following intentionally: Cut myself, burned myself, punched or banged myself, scratched myself, pulled my hair, bit myself, interfered with wound healing, carved words or symbols into skin, punched or banged an object to hurt myself, other?
(5) Suicidal ideation was assessed using the single binary item (yes/no): In the past year, did you ever seriously think about attempting suicide?

## Data Availability

Data are available upon request at https://healthymindsnetwork.org/

